# A Multisite, Randomized Trial Testing a Community-Digital Health Intervention among Black and Latino Adults with Cardiometabolic Conditions: The Roots of Wellness (Raíces del Bienestar) Protocol

**DOI:** 10.64898/2026.05.26.26354175

**Authors:** Cheryl R. Himmelfarb, Joyline Chepkorir, Hailey N. Miller, Oluwabunmi Ogungbe, Nancy Perrin, Wuraola Olawole, Gloria Cain, Ballington L. Kinlock, C. Daniel Mullins, India Kutcherman, Patricia Barger, Manuel Diaz-Ramirez, Jhoselyn Rodriguez, Rosalynn Trujillo, Anna González-Salinas, Roger Clark, Elizabeth L. Andrade

**Affiliations:** School of Nursing, Johns Hopkins University School of Nursing, Baltimore, Maryland; School of Social Work, Howard University, Washington D.C; School of Community Health and Policy, Morgan State University, Baltimore, Maryland; School of Pharmacy, University of Maryland Baltimore, Baltimore, Maryland; Baltimore CONNECT; La Clínica del Pueblo; Coaching Salud Holística; Community Research Advisory Council, Johns Hopkins Institute for Clinical and Translational Research, Baltimore Maryland; Milken Institute School of Public Health, George Washington University, Washington D.C

**Author notes:** Address for correspondence Cheryl R. Himmelfarb 525 N. Wolfe Street, Baltimore, MD, 21205. **Author contribution**: CRH, RC, ELA, HNM, OO and NP conceptualized the study with input from GC, CDM, and BLK. JC wrote the original draft of the manuscript and subsequent revisions with critical feedback from CRH, ELA, HNM, OO, NP, RC, GC, WO, and IK. PB, JR, RT, AG-S, and MD-R contributed to the refinement and finalization of the study protocol and trial implementation. All authors contributed to preparation of the manuscript and have read and approved the final version.

**Keywords:** Black, Latino, hypertension, diabetes, overweight, obese, body mass index, SMS, digital, health promotion, cardiometabolic, cardiovascular, service utilization, health behavior, community health worker, social determinants, trial, community-based

## Abstract

**Background:** Black and Latino adults in the United States experience a disproportionate burden of cardiometabolic conditions due to interacting behavioral, social, and structural drivers of health. Less is known about the impact of integrating digital health tools into CHW-led interventions to improve cardiometabolic health. This trial evaluates a multilevel community–digital health promotion model delivered by CHWs to improve service utilization, health behaviors and cardiometabolic health among Black and Latino adults.

**Methods:** This community-partnered trial uses a randomized delayed-control group with a phased recruitment design. Four cohorts (N = 664) are enrolled through three community-based organizations (CBOs). Eligible participants are ≥18 years who self-identify as Black or Latino, and have prediabetes/diabetes, hypertension, or overweight/obesity. Participants are allocated to either (1) a multilevel intervention consisting of CBO and CHW capacity building combined with individualized CHW-led lifestyle coaching and group activities supported by digital tools, or (2) a delayed control group receiving SMS-only cardiometabolic health education. Data collected at baseline, 6, 9, and 18 months include surveys and health metrics. Qualitative data are collected from participants and community partners to assess intervention acceptability, implementation facilitators and barriers, and sustainability.

**Results:** The primary outcome is health service utilization at 6 and 9 months. Secondary outcomes include health behaviors, health metrics, and social determinants of health. Sustainability of health behaviors and health metrics is assessed at 18 months.

**Conclusions:** Findings will provide evidence to inform scalable, sustainable community-digital health models for CHW-supported cardiometabolic health interventions in underserved communities.

**Trial registration:** Prospectively registered at ClinicalTrials.gov (NCT06607341) on May 23, 2025.

**Sponsor name and contact:** National Institute of Health/National Heart, Lung and Blood Institute 9000 Rockville Pike, Bethesda, Maryland 20892 Phone: (301) 402-9612;

## INTRODUCTION

Cardiometabolic diseases, including diabetes, hypertension, and obesity, affect a large proportion of adults in the United States (U.S.). The vast majority of US adults have, or are at risk for, cardiometabolic risk factors: over 70% are overweight, nearly 42% have obesity, 47% have hypertension, and 57% have diabetes or prediabetes.^1^ These conditions are leading contributors to morbidity, mortality, and healthcare expenditure in the U.S. For example, the economic burden of diagnosed diabetes alone exceeded $400 billion in 2022, reflecting both direct medical costs and lost productivity.^2^ Obesity has similarly been shown to be a major upstream driver of metabolic dysfunction and cardiovascular disease (CVD).^3^ Recent global-burden estimates identify high body-mass index (BMI) as the leading modifiable risk factor for type 2 diabetes and other non-communicable diseases.^4^ Hypertension frequently co-occurs with diabetes and obesity, accelerating vascular complications, renal disease, and premature mortality.^5^

Despite advances in prevention and treatment, substantial cardiometabolic health disparities persist across racial and ethnic groups in the U.S., particularly among Latino and non-Hispanic Black adults.^6^ These disparities are driven by intersecting social and structural determinants such as food insecurity and limited health-care access, which contribute to higher disease prevalence, poorer disease control, and worse cardiometabolic outcomes.^7,8^ Interventions targeting multiple levels of influence (i.e., individual, interpersonal, organizational and community, policy and societal levels) have demonstrated stronger and more sustainable outcomes in chronic, cardiometabolic disease prevention and management than single-level approaches.^1,9^ By simultaneously addressing upstream contextual or structural influences and downstream individual behaviors, multilevel strategies can overcome the limitations of interventions focused solely at one level.^10^ However, few multilevel interventions have been rigorously evaluated in real-world community settings with populations experiencing persistent structural disadvantage, particularly Black and Latino adults at risk for cardiometabolic disease.^11,12^ As a result, empirical guidance remains limited on how to effectively integrate digital tools and community health worker (CHW)-led approaches within existing community infrastructures to promote scalable and sustainable cardiometabolic disease prevention and management.

A robust evidence base supports community-based interventions, particularly those involving CHWs, as effective approaches for improving self-management and clinical outcomes among individuals with cardiometabolic disease.^13,14^ CHWs’ shared lived experience, cultural alignment, and deep knowledge of community resources and social networks, uniquely position them to deliver culturally responsive health and social services within their communities.^15^ For instance, a randomized controlled trial of adult Medicaid beneficiaries in an urban outpatient clinic demonstrated that daily glucose monitoring combined with CHW support produced modest but significant reductions in hemoglobin A1c (HbA1c) among individuals with poor glycemic control, compared with usual care.^14^ Similarly, CHW-led hypertension programs have consistently achieved meaningful blood pressure (BP) reductions.^16,17^ By serving as trusted liaisons between health systems and communities, CHWs help reduce structural barriers to care and promote cardiometabolic disease prevention and management.

Simultaneously, digital health technologies such as SMS/text messaging, telehealth coaching, and web-based platforms have demonstrated strong potential to augment reach, scalability, and engagement in cardiometabolic disease prevention and management.^21,22^ Integration with community-based support, such as CHWs and local organizations, can further strengthen effectiveness, particularly in underserved populations.^15^ Such approaches may be particularly valuable in resource-limited or underserved settings where traditional in-person, clinic-dependent delivery models are constrained.^18,19^ Models that blend in-person CHW contact with digital components are increasingly recognized as sustainable strategies for cardiometabolic disease management.^20,21^ These approaches align with the World Health Organization’s recommendations to develop community-rooted digital health capacity, ensuring long-term sustainability.^22^

Despite the potential to improve reach and impact on health outcomes by using a hybrid approach for cardiometabolic disease prevention and management, few studies have systematically combined CHW-led approaches with digital strategies for multilevel interventions in community-based trials targeting Black and Latino adults with prediabetes/diabetes, hypertension, and overweight/obesity diagnoses. Moreover, little evidence exists on fidelity, sustainability, and long-term outcomes of such interventions. Therefore, the objective of Roots of Wellness (Raíces del Bienestar) is to compare the effect of a multilevel, CHW-delivered, hybrid (in-person + digital) intervention versus an SMS delayed-control intervention among Black and Latino adults with prediabetes/diabetes, hypertension, or overweight/obesity in the District of Columbia (D.C.) and Maryland region. The study aims are:

**Aim 1:** To compare the effect of a multi-level, community-digital health promotion intervention versus delayed control in improving at 6- and 9 months, health service utilization, health behaviors, and health metrics as follows:

1. health service utilization (referral to, utilization of, and satisfaction with),
2. health behaviors (fruit and vegetable intake, sugary beverage intake, exercise, smoking), and
3. health metrics (HbA1c, BP, BMI and waist circumference). **H1.1:** Increase in health service utilization will be greater among participants in the intervention group versus delayed control group at 6 and 9 months. **H1.2:** Improvements in health behaviors will be greater among participants in the intervention group versus delayed control group at 6 and 9 months. **H1.3:** Improvements in health metrics will be greater among participants in the intervention group versus delayed control group at 6 and 9 months.

**Aim 2:** To compare the effect of a multi-level, community-digital health promotion intervention versus delayed control in improving the following determinants at 6 and 9 months: 1) social determinants of health (e.g., housing, food access, digital health literacy, social support), 2) trust in health information sources, and 3) trust in medical research.

**H2.1:** Improvements in social determinants of health, trust in information sources, and trust in medical research will be greater among participants in the intervention group versus delayed control group at 6 and 9 months.

**Sustainability Aim 3:** Among intervention group participants, assess maintenance of the effect of a multi-level, community-digital health promotion intervention on health behaviors and health metrics from 9 to 18 months.

**H3.1:** Participants in the intervention group will maintain improvements in health behaviors and health metrics at 18 months at levels equivalent to or greater than those observed at 9 months.

## METHODS

The Roots of Wellness (Raíces del Bienestar) trial is part of the **C**ommunity **E**ngagement **AL**liance **– D**istrict of Columbia**, M**aryland, and **V**irginia (CEAL–DMV) collaborative. The CEAL-DMV is a collaborative consortium comprising five academic institutions—Johns Hopkins University, George Washington University, Howard University, Morgan State University, and the University of Maryland, Baltimore—and three community-based organizations. Roots of Wellness builds on over five years of partnership and community-engaged research, in which our consortium identified pressing barriers to cardiometabolic health and co-designed solutions with CBOs and community members. This study used a community-based participatory approach with community and academic partners jointly informing all aspects of the trial. This protocol adheres to the Standard Protocol Items: Recommendations for Interventional Trials reporting guidelines. ^23^

### Conceptual framework

Our study is guided by a multilevel conceptual framework integrating the National Institute on Minority Health and Health (NIMHD) Disparities Research Framework and the National Academy of Medicine (NAM) Framework for Assessing Community Engagement in Research **(Figure 1**).^24,25^ These implementation frameworks emphasize how structural, community, interpersonal, and individual determinants shape health disparities and clarify the distinction between upstream systemic drivers and downstream access barriers.^24,25^ Consistent with NAM community engagement principles, community partners participate in design, implementation, and decision-making to ensure cultural relevance and feasibility. This integrated model operationalizes the mechanisms through which CHW-supported strategies enhance health service utilization, social support, and technology engagement, thereby linking individualized needs with community resources to promote health and disease management in minoritized populations. Intervention activities are additionally informed by the Social Cognitive Theory (SCT), which highlights the importance of behavioral capability, self-regulation, and social reinforcement.^26,27^ SCT constructs guide individual-level components such as goal setting, digital skill-building, and coaching, along with social modeling via digital content and group-based lifestyle activities.

**Figure 1:**
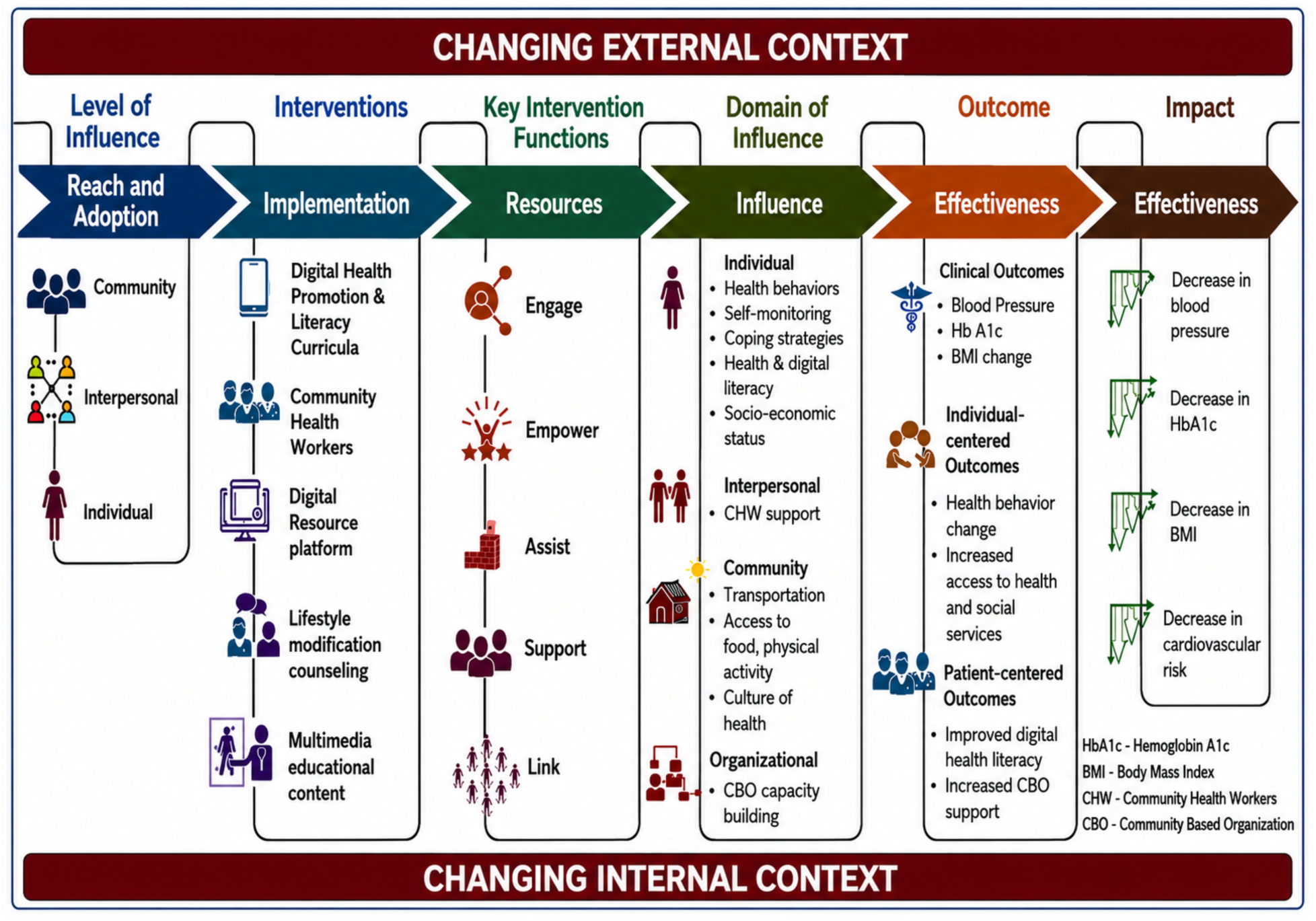
Roots of Wellness Conceptual Framework.

### Setting

Partnerships were established with the following three key CBOs to conceptualize, design, and implement the trial: 1) *Baltimore CONNECT,* based in Baltimore, Maryland, 2) *La Clínica del Pueblo* (LCDP), based in Washington D.C. and Maryland, and 3) *Coaching Salud Holística* (CSH), based in Maryland. Partner CBOs lead participant recruitment and enrollment efforts, hire CHWs and are responsible for implementing and delivering the intervention within their respective communities.

### Design

This trial employs a randomized delayed-control design with a phased recruitment approach. Participants (N=664) are enrolled into four sequential cohorts (A–D), with Cohorts A (Baltimore CONNECT, N=166) and B (CSH, N=83 and LCDP, N=83) recruited in Phase I and Cohorts C (Baltimore CONNECT, N=166) and D (CSH, N=83 and LCDP, N=83) recruited in Phase II (Figure 2). Individuals are randomized in a 1:1 ratio to either the multilevel community–digital health promotion intervention or the delayed-control condition consisting of weekly cardiometabolic disease education delivered via SMS. Delayed-control participants cross over to receive the multilevel intervention at 9 through 18 months, thus ensuring access to all study participants to intervention components while maintaining the ability to evaluate comparative effectiveness. Intervention participants are followed through 18 months to assess maintenance of effects. The trial evaluates 6- and 9-month changes in health service utilization (primary outcome at 9 months) and secondary behavioral outcomes and health metrics (HbA1c, BP, BMI, and waist circumference).

**Figure 2:**
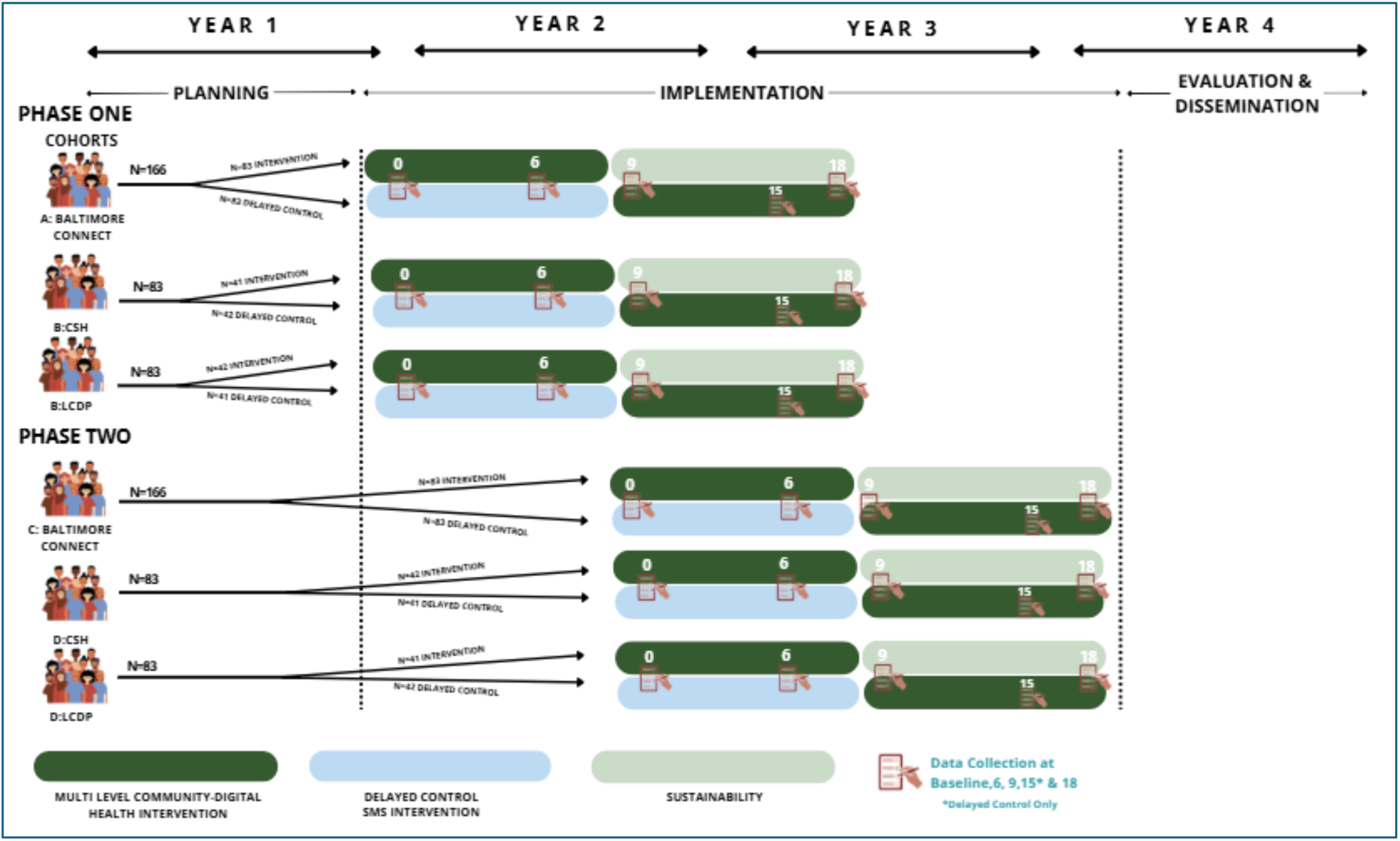
Roots of Wellness Study Design and Timeline.

### Study population and eligibility criteria

This trial recruited includes Black and Latino adults with self-reported diagnosis of prediabetes/diabetes, hypertension, and/or overweight/obesity. The inclusion and exclusion criteria are outlined in Table 1.

**Table 1:** Eligibility Criteria.

| **Inclusion** |
| --- |
| 1. Self-identify as Black or Latino<br>2. Age 18 years or older<br>3. Reside in Maryland or Washington D.C. in high poverty index neighborhoods<br>4. Self-reported diagnosis of one or more target conditions:<br>– Prediabetes/diabetes (HbA1c >5.7%)<br>– Hypertension (BP ≥130/80 mm Hg)<br>– Overweight/obesity (BMI >25 kg/m<sup>2</sup>) |
| **Exclusion** |
| 1. Unable to provide written informed consent in English or Spanish<br>2. No access to a digital device that can connect to the Internet (i.e., laptop, smartphone, tablet or iPad) to allow participation in intervention |

### Recruitment

Eligibility screening and recruitment is conducted by staff of partner CBOs at community events, faith-based institutions, local clinics and other settings routinely accessed by Black and Latino adults. Recruitment for each of the 2 Phases is conducted in an intensive 2-month recruitment period with all participants recruited for a given phase before start of intervention for that Phase. Eligibility screening is completed by CBO staff using a brief survey via Research Electronic Data Capture software (REDCap). Individuals who meet the eligibility criteria are provided with an electronic informed consent form through REDCap or in person, based on participant preference. Participants are considered enrolled in the study once they have completed the baseline survey, including health metrics assessment (HbA1c], BP, BMI, and waist circumference). Participants receive incentives ($50 at baseline, 9- and 18-months and $25 at 6 months) for each completed assessment.

### Randomization

In Phase 1, Cohorts A (Baltimore CONNECT, N=166) and B (CSH, N=83 and LCDP, N=83) are randomized in a 1:1 ratio to either (1) the multilevel community–digital health promotion intervention or (2) the delayed control condition (cardiometabolic health education via SMS). Phase II follows the same randomization procedures for Cohorts C and D. All randomization is computer-generated and stratified by site to ensure balance in geographic and population characteristics across study arms. The allocation sequence is embedded within the REDCap randomization module, which conceals allocation until the moment of assignment. Trained research staff at partner CBOs obtain electronic informed consent and complete baseline assessments before treatment assignment is revealed. Recruiters and CHWs cannot access, view, or modify the allocation sequence at any point.

### Blinding

Data collectors remain blinded to group assignment throughout baseline and follow-up assessments. They are instructed not to inquire about intervention participation and have restricted REDCap access that prevents viewing allocation status.

### Study timeline

The first year of this study involves onboarding community partners, training CHWs and recruiting the Phase 1 cohorts (A and B). The second year includes, for cohorts A and B, the transition of control group participants to the intervention group and sustainability of the intervention participants. Phase 2 begins with recruiting and enrolling the remaining two cohorts (C and D). The final phase of the study involves conducting maintenance assessments and qualitative follow-up interviews with study participants and community partners (CHWs and site leaders). The fourth project year involves primarily evaluation and dissemination activities.

Figure 2 illustrates the study design and phases.

### Trial intervention arms

#### Community-digital health promotion arm

The *Roots of Wellness (Raíces del Bienestar)* community-digital health promotion intervention is delivered at multiple levels, including components at the individual, group, and community levels to reduce risks related to hypertension, overweight/obesity, and (pre)diabetes (See **Table 2**). The intervention was developed collaboratively with community partners, and includes the following components, which are delivered both in-person and digitally: 1) community-digital cardiometabolic health education and CBO capacity building; 2) individualized health coaching to improve lifestyle behaviors and cardiometabolic disease management; 3) individual referrals to social and health services and resources to address social determinant needs using a digital community resource referral platform; and 4) group-based health promotion, including fitness activities and participation in a private Facebook group with weekly cardiometabolic disease prevention content.

**Table 2:**
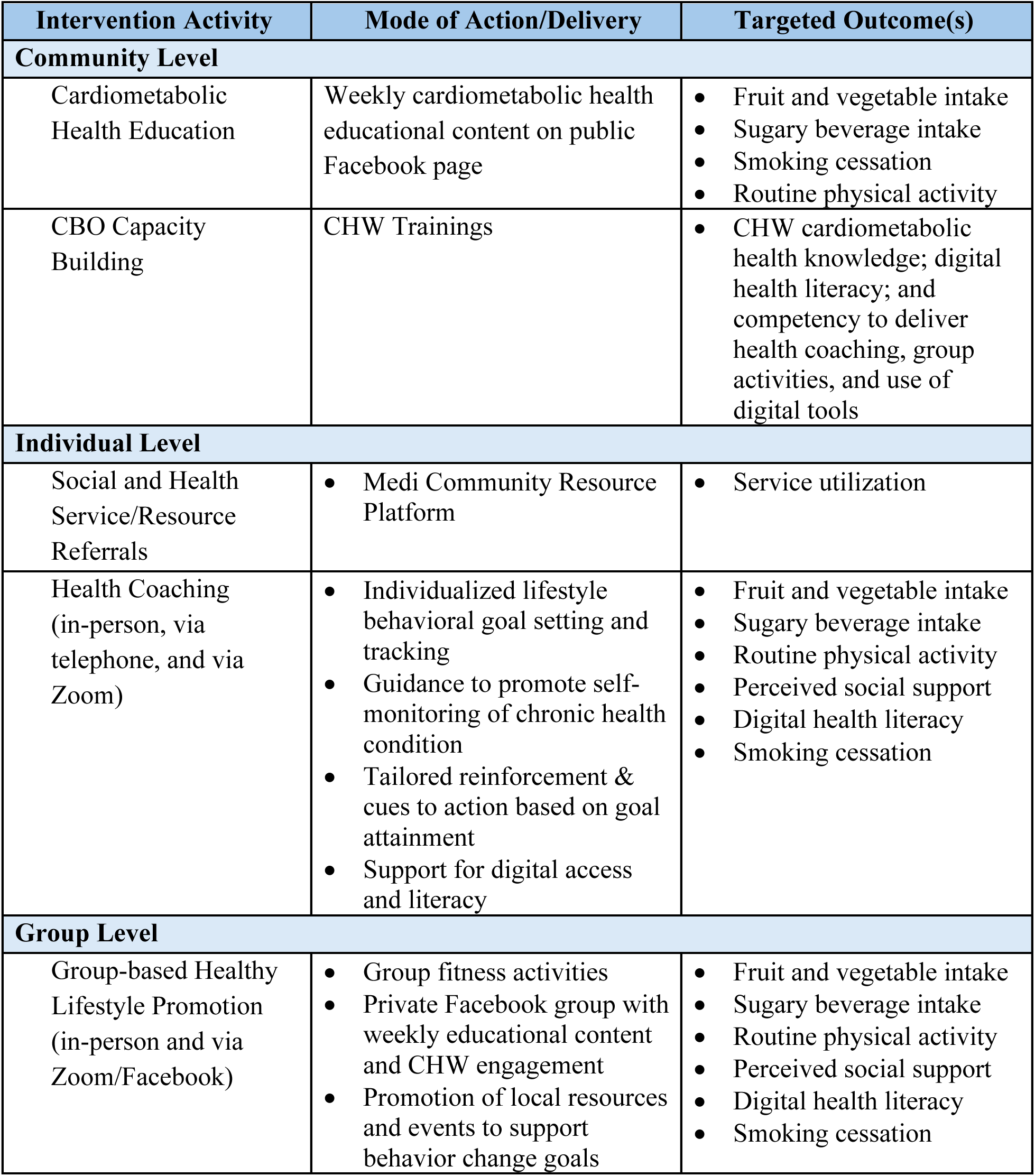
Multilevel community-digital health promotion intervention components.

Health coaching and group-based health promotion address *Life’s Essential 8* (American Heart Association) to guide healthy lifestyle behavior change, such as increasing physical activity, eating a healthy diet, and getting adequate sleep, as well as managing stress and chronic health conditions (i.e., managing blood pressure or HbA1c, taking medications as prescribed, managing weight per medical advice, or adhering to other treatment plans). Intervention components, including individual health coaching, Facebook group-delivered educational content, and group activities, are guided by an adapted 9-month CDC Prevent T2 curriculum (adaptation includes abbreviation of the original 12-month curriculum and addition of hypertension prevention concepts). All intervention activities are delivered by trained CHWs from partner CBOs in English or Spanish, using culturally- and context-appropriate approaches. The CHWs use the Medi (themedi.org) platform to connect individuals to existing healthcare and social services in the community that can support health and behavior change, such as access to food, transportation, housing, financial, and legal assistance as well as dental, medical, mental health and vision care.^28^

CHWs from partner CBOs are trained on intervention delivery using a comprehensive seven-module training curriculum (discussed in detail elsewhere).^29^ The curriculum, co-developed with community partners, covers cardiometabolic disease prevention and management, digital health literacy, using the Medi platform for service referrals, health coaching, digital and group health promotion, and using digital platforms and software for intervention delivery and documentation. Training is conducted over 7 sessions, either in-person or virtually, with each session lasting approximately three hours.

As this is a low-risk behavioral intervention, no formal dose modification criteria apply. Participants may withdraw from the intervention at any time without penalty. The study team may discontinue a participant’s involvement in the intervention in cases of adverse events, inability to adhere to study procedures, or loss to follow-up.

#### SMS delayed control arm

The control group receives weekly cardiometabolic health educational messages via text message in English or Spanish depending on preferred language reported at baseline. The content of the messages encourages core components of *Life’s Essential 8* and includes links to online, reputable resources that provide additional information on the recommendations or education.

Text messages were adapted from the CONNECT study, which developed content in close collaboration with community leaders and cardiometabolic health experts.^30^ All text messages are programmed and sent via Mosio, which is integrated with REDCap.

#### Concomitant care

Participants may be receiving ongoing clinical care, including pharmacologic treatment for diagnosed conditions. The study does not restrict or modify concomitant care. Participants are advised to continue their usual care, and any changes in medications or clinical management are determined by their healthcare providers and are not influenced by the study.

#### Power and sample size calculation

We examined detectable effect sizes for a time-by-group interaction at 9 months (Table 3). Power increased substantially from 250 to 300 participants per arm and more modestly from 300 to 350 per arm. Based on these projections, we recruit a total of 664 participants (332 per arm; 166 per cohort) to allow for an anticipated 10% attrition, yielding 598 participants at 9 months. This sample size provides 80% power to detect an effect size of 0.23 or greater with an α = 0.05 for the primary outcome.

**Table 3:** Effect size detectable for various sample sizes.

| **N per arm** | **Effect Size Detectable** |
| --- | --- |
| 250 | 0.33 |
| 300 | 0.23 |
| 350 | 0.21 |

#### Data collection methods

Data collectors receive standardized training prior to data collection on study procedures and REDCap data entry to ensure consistency and data quality. All study materials (e.g., consent form, surveys, call logs) are professionally translated to Spanish to ensure linguistic and cultural appropriateness. Eligibility screening and assessment of study measures are conducted electronically via REDCap. Data are collected at four time points: baseline (in person), 6 months (phone), 9 months (in person), and 18 months (in person). In-person survey administration takes approximately 60 minutes, whereas phone surveys take about 30–40 minutes. Intervention delivery forms are used to document group activities as well as individual baseline and follow-up assessments completed by CHWs during monthly one-on-one health coaching sessions conducted over the 9-month intervention period.

This study incorporates survey items from the CEAL Common Survey (Cycle 4.0), a standardized instrument developed by the Community Engagement Technical Assistance Center and administered across National Institutes of Health (NIH)-funded Community Engagement Alliance (CEAL) sites. The survey is designed to collect comparable data across regions and to assess the needs, resources, and concerns of communities experiencing health disparities.^34^ Additional items, adapted from validated instruments and supplemented with measures developed by the research team, are also included. **Table 4** provides a detailed list of variables, measures, and frequency of data collection.

**Table 4:** Data Measurement.

| Variable | Measurement | Frequency<br>(in months [M]) |
| --- | --- | --- |
| **Primary outcome** |  |  |
| Health service utilization | Total number of visits (primary care and mental health services)<sup>34</sup> | Baseline (past 6 months); 6M, 9M, 18M |
| **Secondary Outcomes** |  |  |
| **Health behavior** |  |  |
| Dietary intake | Fruits and vegetables (frequency per week)<sup>35</sup><br>Sugary drinks (frequency per week)<sup>35</sup> | Baseline; 6M, 9M, 18M |
| Physical activity | Exercise (in days/week, duration of exercise in minutes)<sup>36</sup> | Baseline; 6M, 9M, 18M |
| Tobacco smoking | Current tobacco smoking status and frequency<sup>37</sup> | Baseline; 6M, 9M, 18M |
| **Health metrics** |  |  |
| Hemoglobin A1C | A1C NOW+ point-of-care device; (normal: HbA1C <5.7%; prediabetes: HbA1C 5.7—6.4%; diabetes: HbA1C ≥6.5%)<sup>38</sup> | Baseline, 9M, 18M |
| Blood pressure (BP) | Calibrated Omron BP monitor (HEM-907XL); average of 3 BP readings obtained with participants seated and a 5-minute rest prior to assessment)<sup>39</sup> | Baseline, 9M, 18M |
| Body mass index | Calibrated Seca portable digital scale and stadiometer (weight in kg/height in m<sup>2</sup>) | Baseline, 9M, 18M |
| Waist circumference | Non-elastic tape measure: circumference measured at the midpoint between the lowest rib and the iliac crest) | Baseline, 9M, 18M |
| **Health-related social needs** |  |  |
| Health-related social needs (housing instability, food insecurity, transportation difficulties, financial strain, utility assistance and employment needs, interpersonal safety, perceived stress) | 12 items adapted from the Accountable Health Communities Health-Related Social Needs Screening Tool (positive/negative screen)<sup>40–45</sup> | Baseline; 6M, 9M, 18M |
| Perceived social and emotional support | Frequency of getting needed social support<sup>46</sup> | Baseline; 6M, 9M, 18M |
| Trust in health researchers and information sources | Perceptions of Research Trustworthiness Scale<sup>47</sup> | Baseline; 6M, 9M, 18M |
| Trust in health information | Trust in sources of information<sup>48</sup> | Baseline; 6M, 9M, 18M |

| **Additional Covariates** |  |  |
| --- | --- | --- |
| Demographic and socio-economic characteristics | Race and ethnicity, gender, age, zip code of residence, education, employment, income<sup>37,49</sup> | Baseline |
| Healthcare access | Health insurance<sup>49</sup> | Baseline; 6M, 9M, 18M |
| Literacy | Single Item Literacy Screener<sup>50</sup> | Baseline; 6M, 9M, 18M |
| Digital health literacy | 10 items adapted from Digital Health Literacy Instrument<sup>51</sup> | Baseline; 6M, 9M, 18M |
| Digital health readiness | 9 items adapted from Digital Health Readiness Questionnaire<sup>52</sup> | Baseline; 6M, 9M, 18M |
| Perceived discrimination in healthcare | Perceived quality of medical care based on racial identity<sup>53</sup> | Baseline; 6M, 9M, 18M |
| Platform specific access | Facebook and Zoom Access | Baseline |

#### Semi-structured interviews

Semi-structured interviews assess perspectives of participants and community partners on the acceptability, barriers, facilitators, and sustainability of the multi-level intervention. Participants are selected using purposeful sampling to ensure representation across demographic characteristics, preferred language, clinical profile, and level of program engagement (utilization of health and social services referred, participation in health coaching and group activities).

Community partners are also invited to provide their perspectives. Each interview lasts approximately 30 to 45 minutes. The semi-structured interview guide is informed by the Consolidated Framework for Implementation Research^51^. Sessions are audio-recorded and professionally transcribed. Participants receive a $50 incentive for their time and contribution.

#### Participant retention and follow-up

Participants receive incentives for completed assessments and are contacted using multiple modalities (e.g., phone, text, and email) to schedule follow-up visits. To promote retention, flexible scheduling and automated reminders are sent via REDCap and SMS. Participants who discontinue or deviate from the intervention will be encouraged to complete all follow-up assessments to minimize missing data.

### Data analysis

#### Primary outcomes

The primary outcome (service utilization at 6 and 9 months) is analyzed using Generalized Estimating Equations (GEE) to estimate population-averaged effects with robust standard errors. Models include time (baseline, 6-, 9-months), group, and group × time interaction, assuming an exchangeable working correlation structure. GEE models yield unbiased estimates under the assumption that data are missing at random. Poisson or negative binominal models are employed depending on the observed distribution of the service utilization variable. Significant interactions are probed using model-based predicted means and graphical displays. Results are reported as effect estimates with 95% CIs and an α=0.05 significance threshold. Adjusted models including baseline variables on which the two groups differ, and variables related to missingness are estimated, if necessary.

#### Secondary outcomes

The same analytical approach (GEE), varying the form of the distribution (normal, logistic, Poisson, negative binomial) depending on the variable of interest, are used to assess the impact of the intervention on health behaviors (e.g. diet and physical activity), health metrics (HbA1c, BP, BMI, and waist circumference), social determinants of health (e.g., housing, food), and trust in medical research and information sources.

#### Missing data

All analyses follow an intention-to-treat approach. The extent and patterns of missing data are examined, and variables associated with missingness identified. GEE approach used provide valid estimates under the assumption that data are missing at random. If the proportion of missing data is substantial, multiple imputation using chained equations are employed. Results from complete-case and imputed analyses are compared to assess robustness.

#### Additional analyses

Sensitivity analyses are conducted to assess the robustness of findings, including models adjusted for baseline imbalances. If adequately powered, exploratory subgroup analyses are conducted based on selected baseline characteristics (e.g., language group or level of intervention engagement). These analyses are considered hypothesis-generating.

#### Sustainability (Intervention arm only)

To assess maintenance of effects from 9 to 18 months, GEE models are used to evaluate within-group change over time among intervention participants only. The delayed-control group is excluded because they initiate the intervention at 9 months.

#### Qualitative data analysis

Data are analyzed using computer-assisted qualitative data analysis software (e.g., NVivo). A multi-coder inductive–deductive approach is employed, with regular consensus meetings to ensure coding consistency. Strategies to enhance rigor include member checking and collaborative interpretation with community partners. Findings from thematic analysis inform ongoing implementation refinement. Community partners contribute to the interpretation of results.

#### Intervention fidelity monitoring

Intervention fidelity is monitored through REDCap-based documentation of all one-to-one coaching sessions, service referrals, and participant engagement in group activities. CHWs complete intervention delivery forms after each participant interaction to capture session content, duration, and participant responsiveness. The study team reviews the use of the Medi by CHWs using monthly reports provided by the Medi executive team. Deviations or implementation challenges are documented and addressed through regular team feedback meetings and retraining as needed.

#### Trial oversight and governance

The coordinating center is located at (institution blinded for review) and is responsible for study design, regulatory oversight, protocol adherence, and coordination across participating sites. A study leadership team, consisting of the Principal Investigators and co-investigators, provides strategic and scientific oversight and meets regularly to review study progress, recruitment, retention, and implementation fidelity. Site investigators and community partners oversee local implementation, including participant engagement and delivery of intervention components. The data management team is responsible for survey design, data management, and quality assurance using REDCap, including routine validation checks and resolution of data queries. Due to the minimal risk associated with this behavioral intervention, a formal Data and Safety Monitoring Board or endpoint adjudication committee was not established. Trial conduct is monitored by the study team, and any issues identified are addressed promptly.

#### Safety monitoring

Given the low-risk, behavioral nature of the intervention, serious adverse events are not anticipated. Harms are defined as any unfavorable or unintended physical, psychological, or social effects experienced by participants during the study period. Adverse events will be assessed through systematic monitoring at data collection points and non-systematically, including participant self-report during intervention activities and assessments. All reported events will be documented, monitored, and reviewed by the study team to determine appropriate follow-up actions.

#### Ethics and dissemination

This clinical trial has received ethical approval from the (institution blinded for review) (IRB# blinded for review), which serves as the single Institutional Review Board (IRB) of record. Participation is voluntary, and informed consent is obtained by trained community partners in participants’ preferred language (English or Spanish) using REDCap. Any protocol modifications are submitted for approval to the IRB prior to implementation. Approved amendments are updated on ClinicalTrials.gov, reflected in study documents and communicated to participants as appropriate. Participant information is collected using secure systems (e.g., REDCap) and stored on password-protected servers with access limited to authorized study personnel. Participants are assigned unique study IDs, and data shared for analysis and dissemination are de-identified. Findings are disseminated through peer-reviewed publications and presentations at scientific conferences. Summary results are shared with participants and community partners in plain language formats, and trial results are reported on ClinicalTrials.gov in accordance with reporting requirements.

### Data availability

De-identified individual participant data (including a data dictionary), statistical code, and relevant study materials will be made available in a publicly accessible repository (e.g., Zenodo, Dryad, or Open Science Framework) following publication of the primary study findings. Data will be shared in accordance with the NIH Data Management and Sharing Policy and institutional guidelines, with appropriate safeguards to protect participant confidentiality.

## DISCUSSION

This study addresses a critical gap in cardiometabolic disease management by evaluating a community-digital health promotion model delivered by CHWs in underserved community settings to improve health and social resource utilization and cardiometabolic health behaviors and outcomes among Black and Latino adults in the DMV region. These populations experience persistent disproportionate cardiometabolic disease burden and current models of care are often inaccessible or do not adequately meet their needs.^52^ Given the continued shortage of healthcare workforce in the US, CHWs are integral to bridging this gap by providing culturally sensitive and linguistically appropriate education (e.g. on Life’s Essential 8) and tangible support (e.g referral to health-related and social resources) that enhance engagement and adherence to health promotion strategies. By leveraging long-standing CEAL–DMV partnerships and community-rooted engagement strategies, the study builds on lessons from prior collaborative efforts, combining grassroots approaches with digital outreach and engagement and access to health and social services for sustainable health improvements. If effective, this intervention is strongly positioned to be integrated into community-based organization infrastructure and health improvement strategies to advance the health of diverse communities.

This trial advances the science of community engaged research in medically underserved Black and Latino communities historically underrepresented in cardiometabolic research.^53^ Previous studies on Black and Hispanic populations have demonstrated the potential of community-engaged strategies to prevent and manage cardiometabolic illnesses.^54,55^ By integrating CHW-led health coaching with low-burden digital modalities (SMS and Facebook), the intervention leverages trusted human support to extend reach, engagement, and scalability.^56^ Phased recruitment further allows the study team to apply lessons learned from earlier cohorts to refine operational processes, strengthen digital components, and adjust CHW workflows and training supports. The randomized delayed-control design preserves internal validity while ensuring all participants ultimately receive the multilevel intervention. ^57^ In particular, the design allows for controlled comparison without indefinitely withholding a potentially beneficial intervention. This was identified by our community partners as essential to building community trust in our team and the Roots of Wellness project.

This study employs participatory research with deep community engagement, a strengths-based transformative approach to build community capacity and improve cardiometabolic health. Community engagement requires meaningfully involving individuals and groups affected by our research goals in the research process.^58^ Using this community engaged approach and formative research, we aim for overall improvement the way the research is prioritized, implemented, translated, and applied in real-world settings, which in turn can lead to improved heath for all.^59–62^ Community-based participatory research has emerged as a transformative implementation research paradigm that bridges the gap between science and practice through community engagement and social action to promote cardiometabolic health.^63^ Indeed, participatory approaches through meaningful community engagement can be instrumental in promoting health by building on the strengths and resources within the community based organizations and the their communities, while promoting co-learning and dissemination of research findings back to the communities;^62,63^ this integration of research knowledge and social action promotes an empowering process (i.e., ‘control over one’s destiny’^64^), which can help develop and implement more acceptable and effective ways to address the health problems faced.

Community partners are engaged in all aspects of Roots of Wellness, starting with identifying key health problems to address and supporting the grant writing process. Key research activities including recruitment and intervention delivery are embedded within partner organizations to enhance trust and build community capacity.^65^ This level of engagement requires deliberate investment by the research team, which is warranted given evidence that community-based organizations, including federally qualified health centers, remain underrepresented in clinical trials due to persistent structural and resource barriers.^66^ The participatory research further supports our implementation science approach involving continuous feedback loops with CHWs and community partners, enabling cultural and contextual adaptation of intervention delivery, while maintaining fidelity to core components.^67^ These iterative processes ensure that the Roots of Wellness model remains responsive to community priorities and supports consistent, high-quality implementation across sites.

Implementation science literature continues to highlight persistent gaps in scalability, sustainability, and context-specific adaptation of health interventions, particularly in underserved settings.^68,69^ Multi-level approaches are required to tackle long-standing, complex health disparities, because these disparities arise from interacting determinants across structural, community, interpersonal, and individual levels.^70^ Building on our integrated conceptual framework, the Roots of Wellness intervention translates these principles into practice through coordinated components that operate across levels, including CHW-led coaching, integration within community-based organizations, and linkage to health and social resources. By aligning intervention strategies with both the NIMHD Research Framework and the NAM Community Engagement Framework, the study moves beyond conceptual application to operational implementation. This distinction is critical, as prior work has demonstrated persistent gaps between theoretical frameworks and their integration into real-world delivery systems, thereby limiting scalability and long-term sustainability of interventions.^71,72^ Furthermore, the incorporation of Social Cognitive Theory strengthens individual-level mechanisms by targeting behavioral capability, self-regulation, and social reinforcement, complementing broader structural and community-level strategies. This multilevel and theory-driven design enhances the potential for sustained behavior change by addressing both proximal behavioral drivers and the contextual constraints that shape them.

### Limitations

The study employs rigorous design and methods, strong community partnerships and established digital outreach workflows. However, several practical and operational challenges may influence implementation outcomes. First, variability in CBO infrastructure and CHW capacity may affect intervention delivery consistency. Site-level heterogeneity arises from differences in staffing, turnover, competing priorities, time for intervention activities, and programmatic orientation. Some sites emphasize group activities or have more stable staffing, and participant profiles vary (i.e., long-standing members regularly engaged in health promotion versus newly recruited participants). Second, digital access barriers such as varying levels of digital literacy may reduce some participants’ engagement with digital group activities including Facebook Group content.^73^

Third, participant engagement and retention is an inherent challenge, particularly in populations experiencing high levels of socioeconomic instability.^74^ Maintaining engagement throughout multiple follow-up cycles requires ongoing coordination among the study team, CHWs, and partner CBOs. To mitigate anticipated attrition, the study incorporates several retention strategies, including collaboration with culturally congruent CBOs and CHWs, proactive communication workflows (e.g., SMS reminders of and follow-up phone calls), and the structured use of incentives to support participant retention.^75^

Finally, although the DMV region’s long-standing academic–community consortium provides an ideal environment for a rigorous implementation trial, this geography-specific structure may limit generalizability. Regions with different infrastructures, CHW workforce models, or policy environments may require adaptation before adopting this model. By explicitly documenting these operational constraints and contextual considerations, the study aims not only to evaluate effectiveness but also to generate actionable guidance for real-world adoption of the intervention.

### Conclusion

This trial evaluates a community-driven CHW-led digital health promotion model for cardiometabolic risk reduction in Black and Latino communities. By linking implementation approach and outcomes to community priorities, we aim to inform scalable, sustainable approaches for advancing cardiometabolic health in diverse populations.

## Data Availability

Not applicable, this is a protocol paper.

## Acknowledgments

We are grateful to the research participants, community partners, community health workers, and data collectors for their contributions to the Roots of Wellness Study.

